# Probabilistic reconstruction of measles transmission clusters from routinely collected surveillance data

**DOI:** 10.1101/2020.02.13.20020891

**Authors:** Alexis Robert, Adam J. Kucharski, Paul A. Gastanaduy, Prabasaj Paul, Sebastian Funk

## Abstract

Pockets of susceptibility resulting from spatial or social heterogeneity in vaccine coverage can drive measles outbreaks, as cases imported into such pockets are likely to cause further transmission and lead to large transmission clusters. Characterising the dynamics of transmission is essential for identifying which individuals and regions might be most at risk.

As data from detailed contact tracing investigations are not available in many settings, we developed a R package called *o2geosocial* to reconstruct the transmission clusters and the importation status of the cases from their age, location, genotype, and onset date.

We compared our inferred cluster size distributions to 737 transmission clusters identified through detailed contact-tracing in the United States between 2001 and 2016. We were able to reconstruct the importation status of the cases and found good agreement between the inferred and reference clusters. The results were improved when the contact-tracing investigations were used to set the importation status before running the model.

Spatial heterogeneity in vaccine coverage is difficult to measure directly. Our approach was able to highlight areas with potential for local transmission using a minimal number of variables and could be applied to assess the intensity of ongoing transmission in a region.

## Introduction

Establishing who infected whom during an outbreak can help inform the design and evaluation of control measures[1-5]. Transmission links can be reconstructed through contact tracing investigation, whereby cases are asked their movements and contacts during their infectious period. Given that contact-tracing investigations are not always carried out due to the logistical effort and cost involved, inference methods have been developed to use epidemiological data to estimate the probability that a transmission event occurred between any given pair of cases[6-12]. This makes it possible to establish probabilistic transmission trees that link all observed cases. The ensemble of cases belonging to the same transmission tree is called a transmission cluster.

Wallinga and Teunis first developed a likelihood-based estimation procedure to reconstruct probabilistic transmission trees from a given distribution of generation times and observed symptom onset dates of each case[2]. Since then, genomic, spatial or contact data have been used to supplement the timing of symptoms, which helped identify determinants of transmission, mixing behaviour, individual dispersion, evaluate control measures, anticipate future developments of outbreaks and study viral evolutionary patterns[5,8,9,13-17].

As sequencing of pathogens has become more common, the use of such data to infer transmission trees has increased. Methods developed to add genetic distance to a Wallinga-Teunis algorithm, where cases with lower genetic distance are more likely to be grouped in the same transmission group, showed it substantially increased the accuracy of the reconstructed transmission trees[8,18-21].

The utility of sequence data depends on the characteristics of the pathogen[22,23]. Based on the highly variable 450 nucleotides region of the N gene (N-450) of the measles virus genome, eight measles genotypes have been detected since 2009[24,25]; these genotype designations are helpful in linking cases, as linked cases must be infected by virus of the same genotype[25]; however, the diversity of measles genotypes is decreasing[26]. It has been suggested that further sequencing the M-F non-coding region, or full genome sequencing, could help identify measles virus transmission trees, but so far, extended sequencing during measles outbreaks has been scarce [27,28]. In addition, the evolutionary rate of measles virus is very low[29], therefore, samples from unrelated cases can be very close genetically and genetic sequences from measles cases are not usually indicative of direct transmission links[27,28].

As measles is highly infectious, under-immunized communities (also called pockets of susceptibles) resulting from local heterogeneity in vaccine coverage can lead to large, long-lasting outbreaks[30- 34]. Detecting these pockets of susceptibles can be challenging, as historical local values of coverage throughout a given country are rarely available. The number of cases in the transmission trees resulting from each importation during outbreaks, also called the cluster size distribution, will depend both on individual factors (e.g. age of the imported case which might affect contact patterns) and community factors (e.g. the history of coverage in the area)[35,36]. The size of a cluster can therefore reflect the level of susceptibility of individuals directly and indirectly connected to the imported case [37,38].

Here we introduced a model combining age, location, genotype, and rash onset date of cases to reconstruct probabilistic transmission trees. We chose these features to make the model applicable to a wide range of settings as they are commonly reported and informative on transmission. We wrote the R package *o2geosocial* to conduct inference on individual-level data using this model. It is based on the package *outbreaker2* and is designed for outbreaks with partial sampling of cases, or uninformative genetic sequences, such as measles outbreaks[9,39]. We used the likelihood of transmission links between different cases to estimate their importation status. We compared the inferred importation status and cluster size distribution to the transmission clusters identified via contact tracing during measles outbreaks in the United States between 2001 and 2016.

## Methods

### Presentation of the algorithm

Transmission trees are used to represent who infected whom during an outbreak. They are directed acyclic graphs, where nodes are the reported cases and edges show the connection between them. The root of each transmission tree is an imported case, i.e. a case who was infected in a different transmission setting. The cases placed in the same transmission tree form a transmission cluster. We estimated the number of cases per cluster (cluster size distribution) and the importation status of the cases from probabilistic transmission trees inferred using routinely collected epidemiological variables.

We used a Metropolis Hastings algorithm with Markov chain Monte Carlo (MCMC) to classify a set of cases into a set of transmission trees with associated probabilities quantified using a Bayesian model to combine the epidemiological features of the cases. At every iteration of the MCMC algorithm, we proposed a new set of model parameters, infection dates and connections between cases. These three elements formed a tree proposal. We computed the ratio between the posterior probability of this proposal and the current posterior probability. The posterior probability (up to a multiplicative constant which would cancel out when calculating the ratio) was calculated from the likelihood of the trees, and the prior probability of the parameters. The log-likelihood of each tree was equal to the sum of the log-likelihoods of each case. All the notations were defined in Table 1.

**Table 1:**
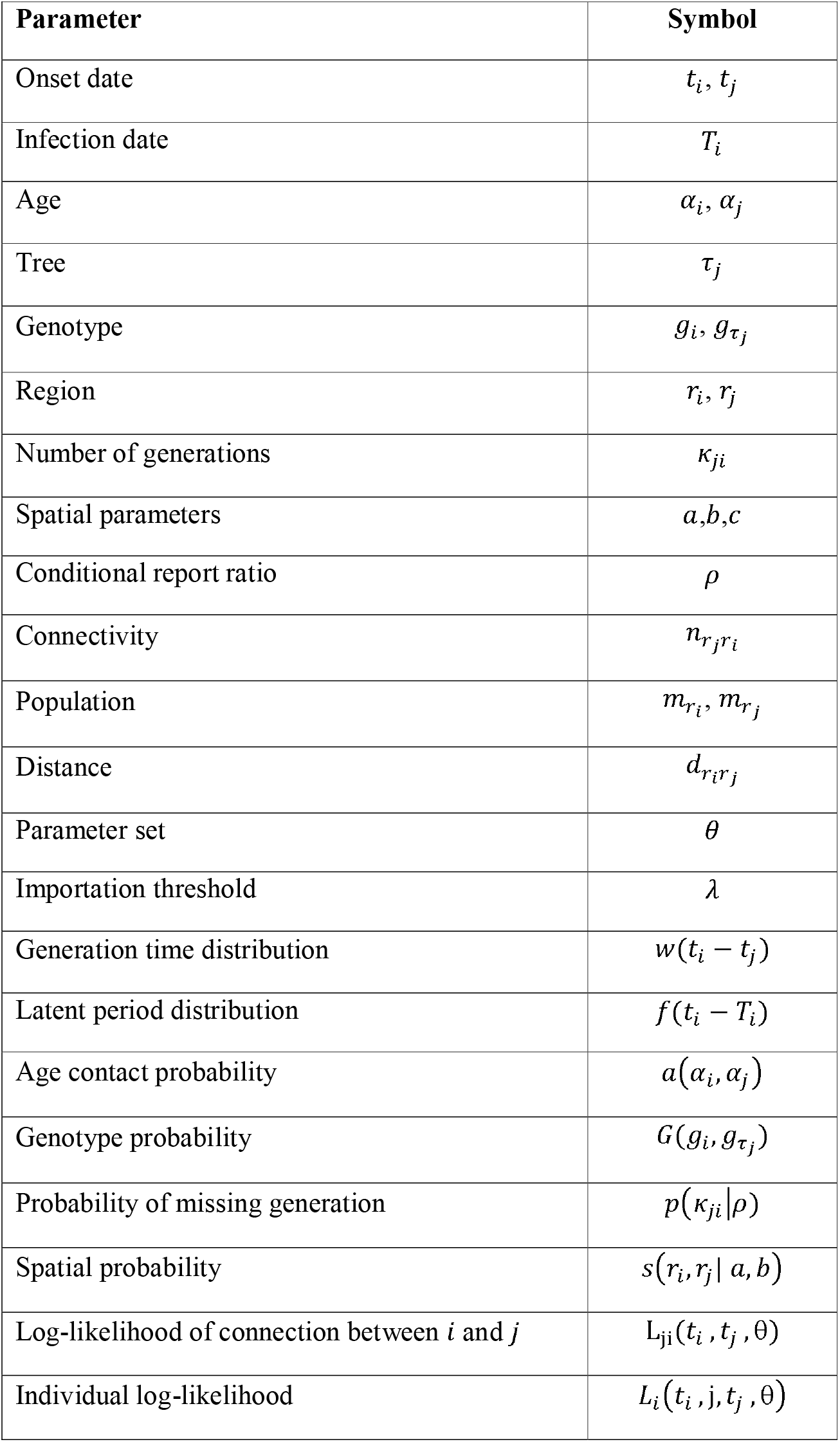
Table of notations of all variables and distributions defined in the methods.

### Likelihood function and parameter definition

In a tree proposal, each case *i* was assigned an infector *j* and an infection date *t_i_*. We computed the log-likelihood of each case, *L_i_*(*t_i_*, *j*, *t_j_, θ)* to calculate the likelihood of the tree. The log-likelihood of *i* was split in two: *i*) the log-probability density of observing the onset date *T_i_* if case *i* had been infected at time *t_i_* log(ƒ(*t_i_* − *T_i_*)); and ii) the log-likelihood of connection between *i* and *j* L*_ji_*(*t_i_*, *t_j_*, *θ*), with *θ* the parameter set of the model (1):

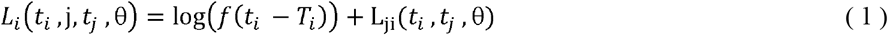

The function *f* represents the distribution of the incubation period. The log-likelihood of connection L_ji_ was computed from five components reflecting the age group, genotype, location, inferred date of infection of cases *i* and *j*, and the report ratio (2). We allowed for undirect link between cases due to unreported individuals, *k_ji_* corresponds to the number of generations between *i* and *j*. If *k_ji_* = 1, *j*, infected *i*, whereas if *k_ji_* = 2, an unreported case infected by *j* infected *i*, *k_ji_* increases with the number of missing links between *i* and *j*.

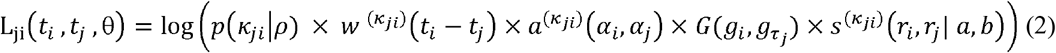

We calculated the temporal probability of transmission between *i* and *j* from the number of days between and *t_i_* and *t_j_* the distribution of the generation time of the disease *w*(t). This probability was quantified by 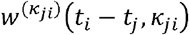, 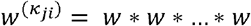, where * is the convolution operator applied *k_ji_* times. We used a geometric distribution 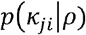 to quantify the probability of observing *k_ji_* missing generation between *i* and *j* given the conditional report ratio *ρ*. The conditional report ratio quantifies the probability of missing generation between two connected reported cases. Entire missing clusters, cases infected after the last cases or cases infected before the ancestor of a cluster would not interfere in the connection between two cases, and therefore would not affect the value of the conditional report ratio. The conditional report ratio can be higher than the overall report ratio of an outbreak. The “ancestor” is the earliest identified case in a cluster.

*a*(*a_i_, a_j_, k_ji_*) was defined as the probability of transmission between age groups *α_i_* and *α_j_*. This probability corresponds to the proportion of contacts to the age group *α_i_* that originated from *α_j_* and can be deduced from studies such as Polymod[36]. We defined 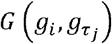 as the probability of observing the pathogen genotype *g_i_* in case *i* in the tree *τ_i_* containing case *j*. There can only be one measles virus genotype per transmission tree, or cases with unreported genotype. The genotype 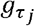 is the genotype contained in the tree *τ_i_* and is known if at least one case in *τ_j_* had a reported genotype.

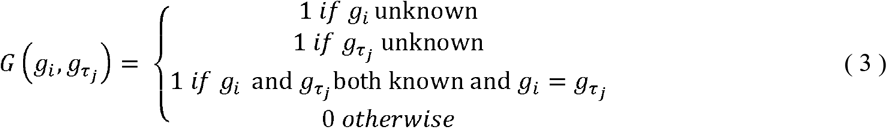

In (3), if 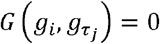, then the connection between *i* and *j* is impossible, and (1) and (2) are equal to *log*(0)= −∞.

*s*(*r_i_, r_j_, k_ij_)* was defined as the probability of connection from *r_j_* to *r_i_* regions of residency of *i* and *j* (4). We used an exponential gravity model to quantify the connectivity of the different geographical units[40]. This approach showed good performance at modelling short distance commuting, and was easy to parametrise[40-44]. In the simplest form of the exponential gravity model, the number of connections between *r_i_* and *r_j_* is proportional to the product of the origin population 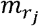, the destination population 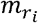. and an exponential decrease of the distance between *r_i_* and 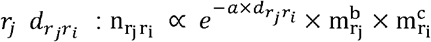, with *a, b*, and *c* parameters adjusting for the impact of distance and population. From this definition, we deduced *s*(*r_i_*, *r_j_*), the spatial probability of transmission from *i* to *j*:

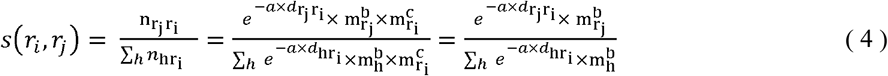

Only the parameters *a* and *b* were required to compute the spatial probability of transmission. If *r_i_ = r_j_*, then (4) becomes: 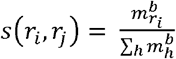, Other distributions than the exponential decrease can be used in this framework if transmission follows a different pattern.

The parameters *p, a*, and *b* were estimated. At each iteration of the MCMC, the log-likelihood of the trees was equal to the sum of all individual log-likelihoods *L_i_* from equation (1). The log-posterior density of the proposed trees was calculated by summing the overall log-likelihood of the trees and the log-priors of the parameters.

### Tree proposals

We used a Metropolis Hastings algorithm with MCMC to sample from the posterior distribution of parameters and the transmission trees. To do this, we developed a set of proposal tree updates. These updates were accepted with acceptance probability as defined by the Metropolis-Hastings algorithm[45]. We used eight types of tree proposal to ensure good mixing. Each proposal conserved the overall number of trees, with a maximum of one unique genotype reported per tree.

Five of the proposals had already been implemented in the *outbreaker2* package and were adapted to this setting: i) change the number of generations between two cases; ii) change the conditional report ratio *ρ*; iii) change the time of infection; iv) change the infector of a case (if the case is not the ancestor of a tree); v) swap infector-infectee (if none is the ancestor of a tree).

We added two proposals to change *a* and *b* the spatial kernel parameters. For each proposal, the probability of transmission between every geographical unit was re calculated with the new values. The distance matrix had to be computed for each number of generations between cases, which considerably slowed down the algorithm. As we could not use sequence data, assessing whether a case was isolated or whether it was connected to a reported infector with two missing generations would be very challenging using our model alone. Therefore, we limited the maximal number of missing generations to 1 when *a* or *b* were estimated (max(*k_ji_*) = 2). Finally, the last proposal was designed to change the ancestor of the tree whilst conserving the overall number of trees (Figure 1).

**Figure 1:**
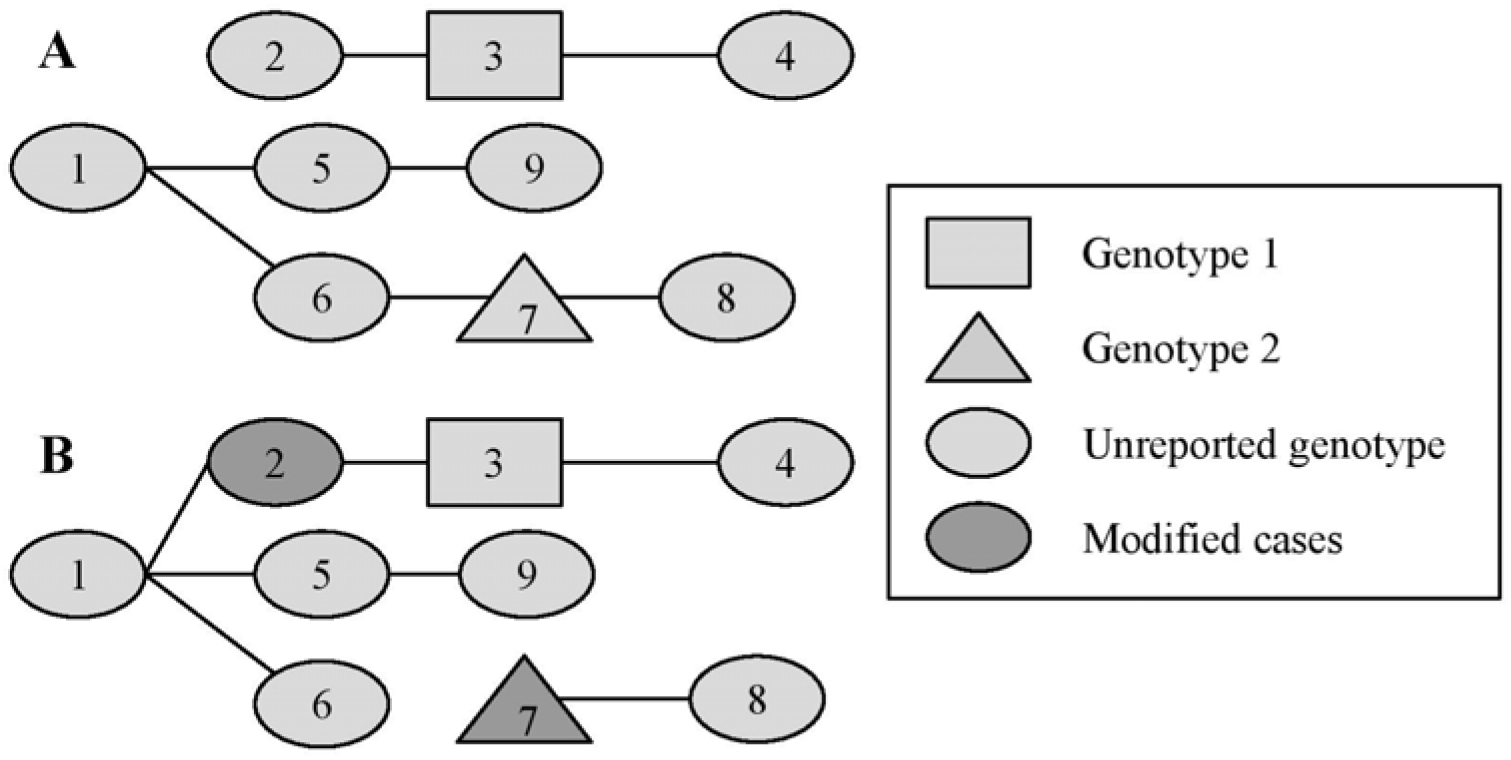
Example of the change of ancestors. Panel A represent the initial tree, B is the new tree proposed after the movement. Initially, there are two ancestors (cases 1 and 2) in a group of 9 cases. 3 and 7 have different genotypes and cannot be part of the same tree, the genotypes of the other cases are not reported. The date of infection is in increasing order (1 is the first case, 9 is the last). Therefore, 1 is the only potential infector for 2. One new ancestor was randomly drawn to conserve the number of trees. In this example, 7 is the new ancestor (6 was the only other possibility). The ratio of the posterior densities of A and B were then used to determine whether to accept or reject the proposal, according to the Metropolis-Hastings algorithm. This movement ensures good mixing of the potential ancestors of the transmission clusters.

### Inference of importation status and cluster

Unrelated measles cases stemming from different importations and different regions can be part of the same dataset. Grouping cases and excluding unrealistic transmission links reduces the number of possible trees and speeds up the MCMC runs. To do so, we listed each case’s potential infectors using three criteria: i) The potential infectors must be of the same genotype as the case, or have unreported genotype, ii) The location of potential infectors must be less than *γ* km away from the case and iii) the potential infectors must have been reported later than δ days before the case. This threshold should be determined from the maximum plausible generation time of the disease. The spatial threshold *γ* should be defined according to the relevance of long-distance transmissions. Cases with no potential infector were considered as importations. Otherwise, they were grouped together with i) their potential infectors and ii) cases with common potential infectors.

After grouping the cases, we estimated their importation status and the cluster size distribution using two runs of MCMC (Figure 2). The first run was shorter and aimed at removing the most unlikely connections among each group, as they can reflect unrealistic estimates for incubation periods or generation times and corrupt the estimation of the date of infection. We defined a reference threshold *λ*, whereby if the individual value of log-likelihood L*_i_* was worse than *λ*, then the connection between *i* and their infector was considered unlikely. In *Outbreaker2, λ* was a relative value, defined from a quantile of the individual log-likelihoods. In *o2geosocial, λ* can be a relative value or an absolute value, chosen from the number of components of the likelihood. For each sample saved from the short run, we computed the number of unlikely connections *n*. If there was no iteration where all connections were better than *λ*, *min(n)* new importations were added to the initial tree for the long run (Figure 2).

**Figure 2:**
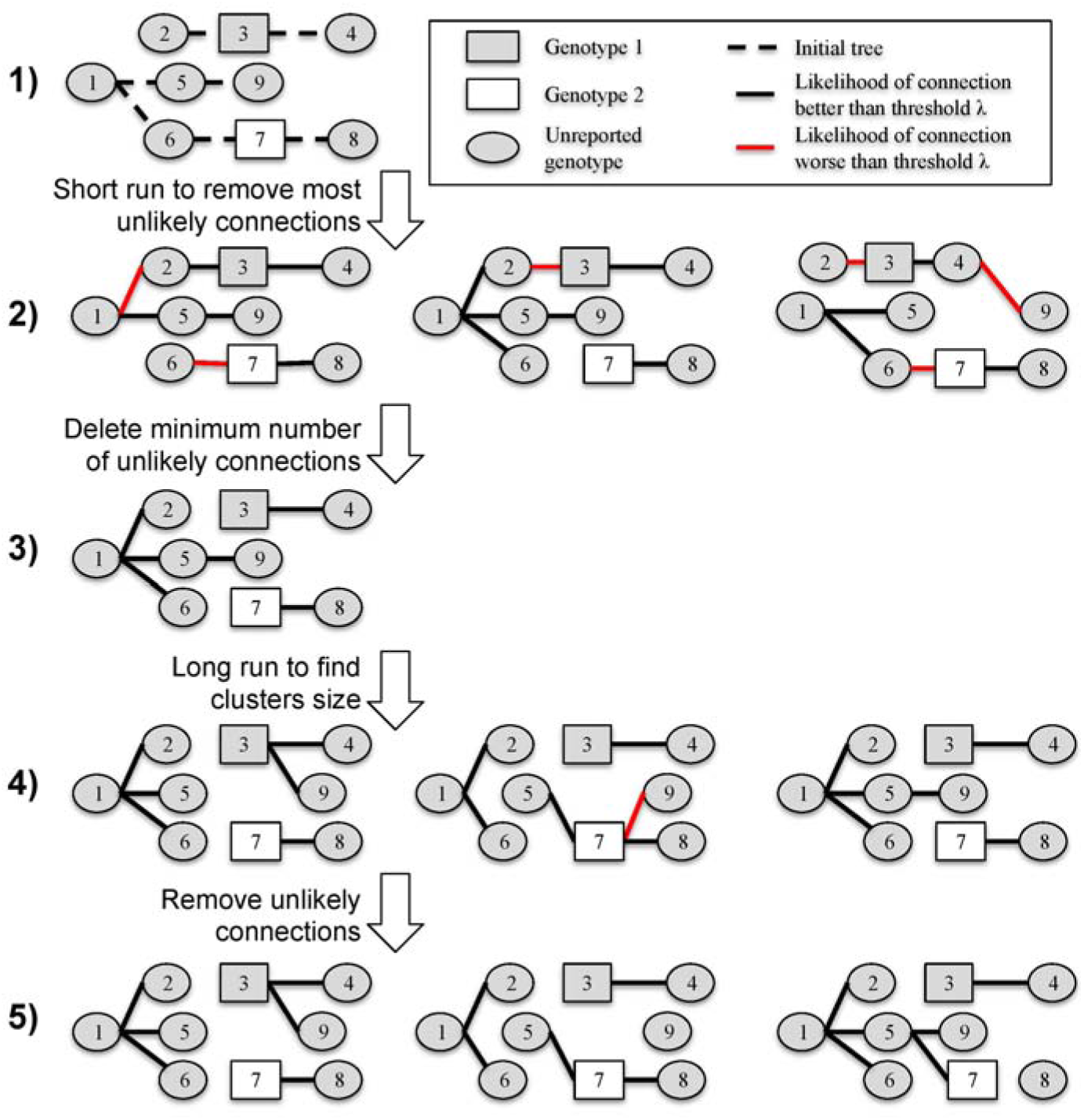
Estimating importation status and cluster size distributions in two MCMC runs. Step 1: Initial tree obtained after pre-clustering, with the minimum number of importations (here 2, as there are two reported genotypes). Step 2: Samples from the first short run, with red line showing connection worse than the arbitrary threshold *γ*. Step 3: Initial tree for the final run, with 1 more importation than in step 1, which corresponds to the minimum number of unlikely transmissions at step 2. Step 4: Samples from the long run. Step 5: Final trees used to compute cluster size distribution and importation status of each case. Case 7 is an importation in one third of the final samples, whereas case 3 is an importation in all of them.

Finally, we ran a long MCMC chain and obtained samples from the posterior distribution. After removing the burn-in period and thinning the chain, we deleted the unlikely transmission links in each iteration and identified transmission clusters. Therefore, unlike the previous versions of *outbreaker2*, the number of importations in each sample can vary and the individual probability of being an importation can be computed (Figure 2).

## Validation case study: measles outbreaks in the United States between 2001 and 2016

### Data

To evaluate the performance of the model, we inferred the transmission clusters from a dataset that also included information on whether measles cases were part of a cluster based on contact tracing investigations. Measles cases in the United States are reported by healthcare providers and clinical laboratories to their corresponding health department. Each case is investigated by local and state health departments classified according to standard case definitions[46], and linked into clusters epidemiologically (e.g., by establishing a direct contact or a shared location between cases, or when cases are part of a specific community where an outbreak is occurring). Cases are considered internationally imported if at least part of the exposure period (7-21 days before rash onset) occurred outside the United States and rash occurred within 21 days of entry into the United States, with no known exposure to measles in the United States during the exposure period.

Confirmed measles cases are routinely reported by state health departments to the CDC. 2,098 measles cases were reported in the United States between January 2001 and December 2016. The number of annual cases did not exceed 700 cases during this time period (Figure 3, Supplement Figure S1). The importation status, 5-year age group, onset date, county, and state of residence were fully reported for 2,077 cases. The 21 cases with missing data were discarded. 25% of the cases were classified as importations. 39% of the cases had their genotype reported.

**Figure 3:**
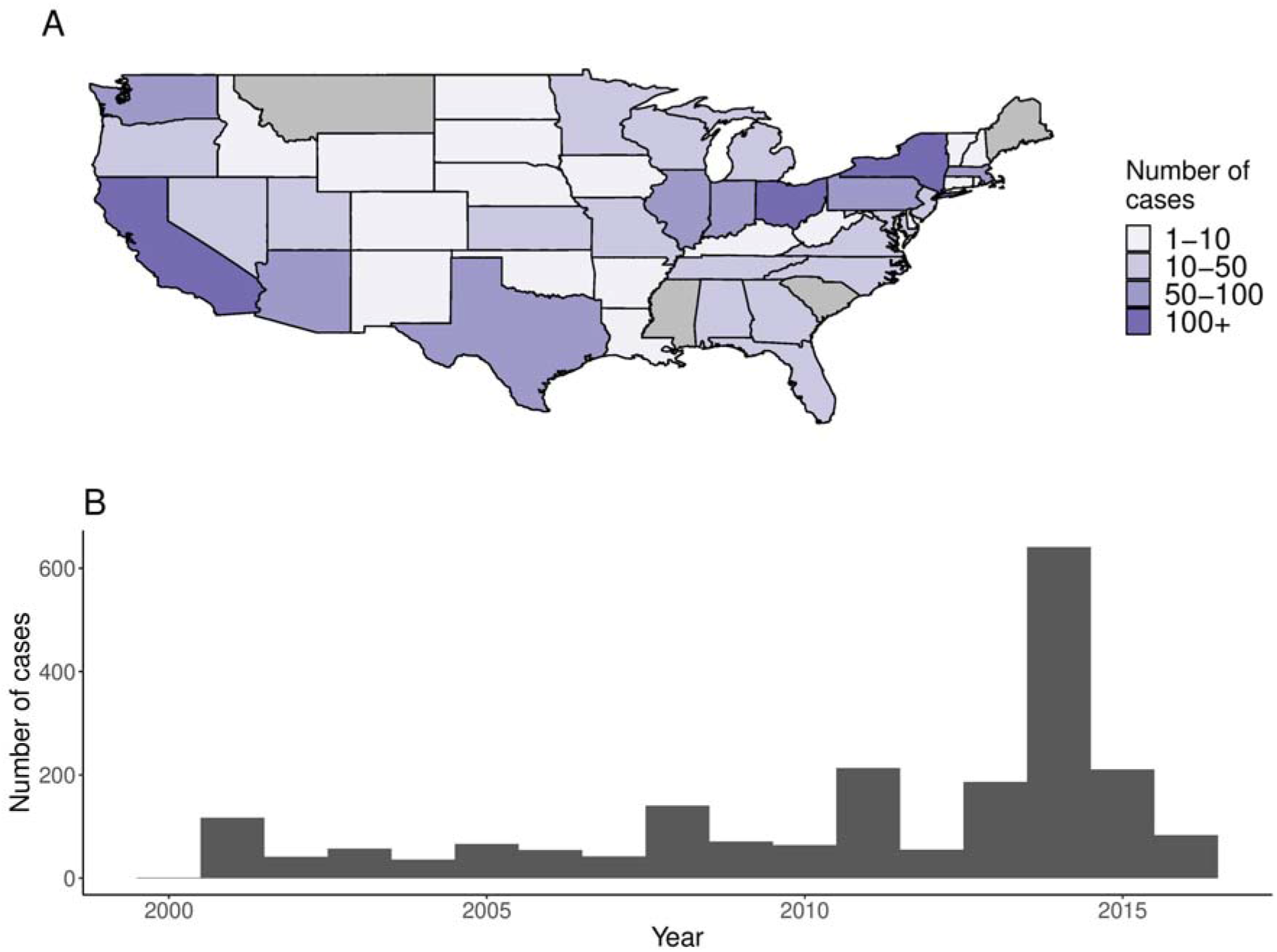
Panel A: number of cases per state and Panel B: Annual number of cases reported in the United States between 2001 and 2016. Alaska and Hawaii are not shown on Panel A.

Among cases with complete data, 737 independent clusters, containing 1 to 380 cases, were reconstructed through contact tracing investigations. Not every identified case could be linked to an importation, and some transmission clusters contained multiple imported cases (e.g. when related individuals travel together to a foreign country and were infected there). Out of the 737 reference clusters, 38 had several cases classified as importations, 256 had none identified.

### Model and parameters

The distributions and priors used in the studies are listed in Table 2. As no studies quantifying the probability of age-specific contacts have been carried out in the United States, we used the estimates from the POLYMOD study in the UK[36]. The incubation period and the generation time of measles were taken from previous studies [47-49]. We used the population centroid of each county to compute the distance matrix[50]. We used a beta distribution as the prior of the conditional report ratio[8]. The mean of the prior distribution was calculated using the number of clusters whose first case was not classified as an imported case, meaning the investigations were not able to trace back to the first case imported. As there was no prior information on the possible values of the spatial parameters *a* and *b*, we used uniform distributions between 0 and 5.

**Table 2:** Values of parameters used to cluster cases declared in the United States.

| Parameter | Symbol | Distribution |
| --- | --- | --- |
| Incubation period | $f(t)$ | Gamma,<br>mean = 11.5, sd = 2.24 |
| Generation time | $w(t)$ | Normal,<br>Mean = 11.7, sd = 2.0 |
| Conditional report ratio | $\rho$ | Prior: Beta distribution,<br>Mean = 0.65, sd = 0.15 |
| Spatial parameter 1 | $a$ | Prior: Uniform distribution |
| Spatial parameter 2 | $b$ | Prior: Uniform distribution |
| Spatial pre clustering | $\gamma$ | Fixed: 100 km |
| Temporal pre clustering | $\delta$ | Fixed: 30 days |
| Importation threshold | $\lambda$ | Absolute:<br>$5 \times \log 0.05 = -15$<br>$5 \times \log 0.1 = -11$<br>Relative:<br>5% |

For pre-clustering of cases, we set the temporal threshold *δ* to 30 days, which is above the 97.5% upper quantile of the generation time with a missing generation. We were interested in local transmission to describe the impact of an imported case on a community. But we only had information on the county of residency for each case. Counties are large geographical units: the average county land area is 2,911km^2^ and the maximum values reach 50,000km^2^. Therefore, we set the spatial threshold *γ* to 100km to exclude long distance transmission, while still allowing for cross-county transmission.

Finally, we tested several relative and absolute importation thresholds *λ*. Absolute values were calculated from a factor *k*, multiplied by the number of components in *L_i_*, excluding the binary genetic component. Tested values were *k = 0.05* (*λ =* log(0.05) * 5 = −15) and *k = 0.1* (*λ* = −11). Connections were considered unlikely if the log-likelihood was worse than *λ*. Relative values were quantiles of all recorded log-likelihoods in the sampled trees (Table 2).

### Inference of importation status

Using the contact tracing investigations, we considered three different initial distributions of the importation status. In scenario 1, there was no inference of the importation status of cases, and the first case of each epidemiological cluster was classified as importation (Ideal importation). In scenario 2: there was no inference of the importation status of cases, and all cases identified as importation in the contact tracing investigations were classified as importations (Epidemiological importation). Finally, in Scenario 3, the importation status of cases was inferred, using different thresholds *λ*, and using no prior information on the importation status of cases or the importation status from the contact tracing investigations.

### Inference of clusters

In order to compare the inferred and reference clusters, we calculated for each case *i*:i) the proportion of cases from the same reference cluster as *i* that were inferred with *i* (sensitivity) and ii) the proportion of cases in the same inferred cluster as *i* that were part of the reference cluster (precision). These values were calculated at every iteration, and the median values were used to evaluate the fit obtained with different values of *λ*. We also used the inferred cluster size distribution to the reference data. The credibility intervals for each case are reported in the Supplement (Supplement Figure S2).

## Results

We clustered 2,077 measles cases reported in the United States between January 2001 and December 2016 using their onset date, age groups, location and genotype. Using the contact tracing investigations, we considered three different initial importation status distribution: i) only the ancestors of each epidemiological cluster (first case of each cluster) were importations (ideal importation), ii) all cases classified as importation in the contact tracing investigations were importations (epidemiological importation), iii) no prior information on importation status of cases. The importation status of the cases was therefore not probabilistically inferred in scenario 1 and 2. The short preliminary run was 30,000 iterations and 70,000 iterations. For each run, the trace of the posterior distribution shows the convergence of the algorithm (Supplement Figure S3).

In scenario 1, we did not infer the importation status of cases. The inferred cluster size distribution matched the contact tracing investigations (Figure 4A); 98% of the reference singletons were also isolated in the inferred cluster. For 94% (95% Credibility Interval: 91-98%) of cases, the inferred cluster had a sensitivity and precision above 75%, meaning more than 75% of the cases in the inferred cluster were in the reference cluster, and more than 75% of the cases in the reference cluster were in the inferred cluster (Figure 4B). For 80% (78 - 93%) of cases, the inferred clusters were a perfect match with the reference clusters. The cluster size distribution stratified by state was similar to the contact tracing investigations (Supplement Figure S4). Therefore, when each ancestor was considered as an importation, the inferred clusters were very close to the reference ones.

**Figure 4:**
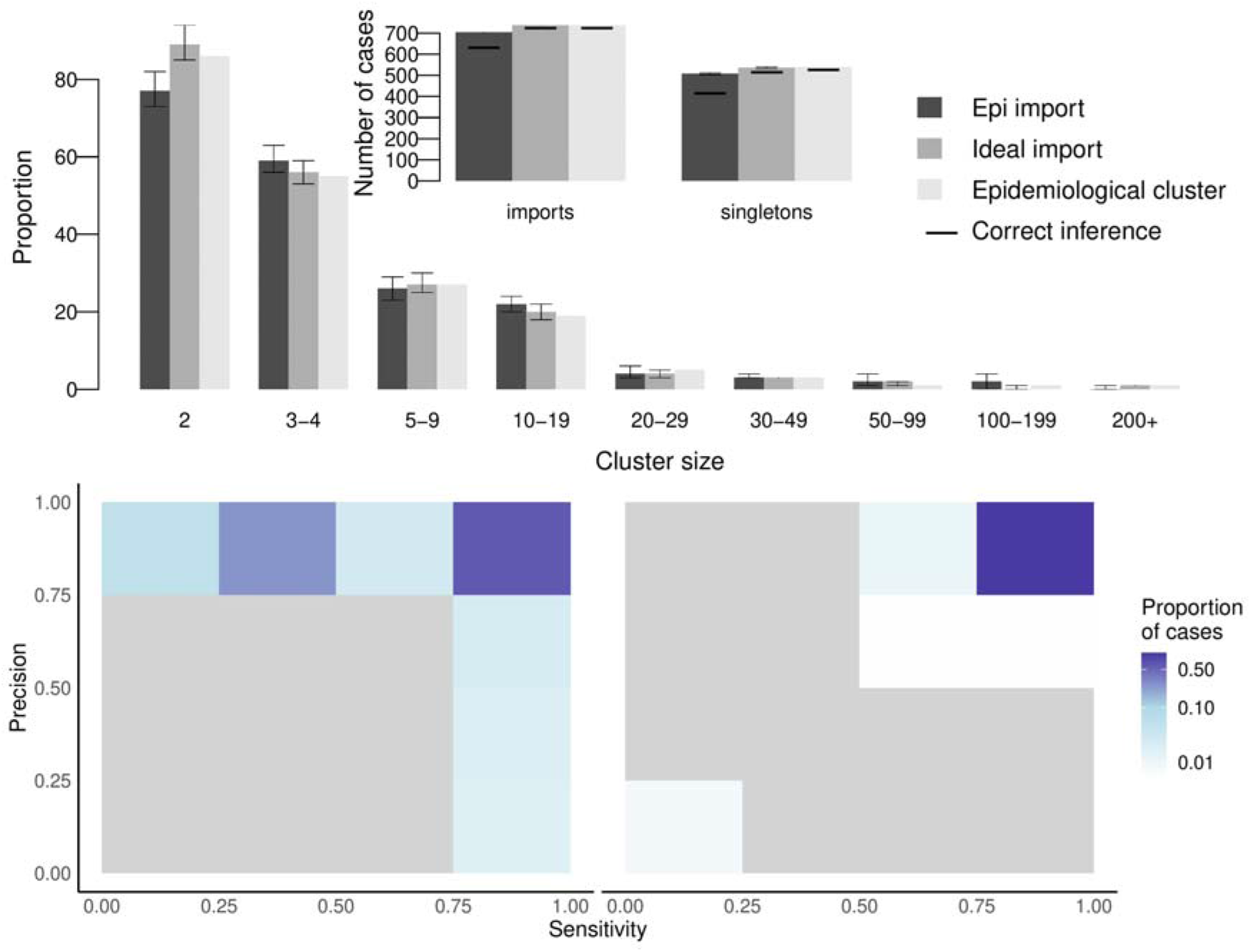
Description of transmission clusters inferred using prior knowledge on importation status of cases. Panel A: Cluster size distribution for the scenario 1 and 2 (grey and dark grey), compared to the reference clusters (light grey). Arrows represent the 95% credibility intervals of each estimate. Only clusters containing at least 2 cases are represented. Insert: Number of importations and number of isolated cases (singletons) in scenario 1 and 2, and in the reference clusters. For each scenario, the horizontal dark line represents the number of importations that are also importations in the reference clusters, same for singletons. Panel B: Heatmap representing the precision and sensitivity of the clusters for each case in scenario 1, cases are classified in a category depending on the proportion of their reference cluster that were inferred in the same cluster (x-axis) and the proportion of mismatches in the inferred cluster. Panel C: Same for scenario 2.

In scenario 2, we used the importation status distribution of cases reported in the contact tracing investigations (539 importations). Pre-clustering highlighted 165 cases with no potential infector, which were also classified as importations. We observed discrepancies between the inferred cluster size distribution and the reference one: Among the 704 cases inferred as importation, 61 (9%) were not importations in the reference cluster. Furthermore, 94 cases were the ancestor of a reference cluster and were not classified as importations in the inferred clusters (13%). The overall cluster size distribution matched the reference distribution, but 111 reference singletons were inferred as part of transmission clusters (Figure 4A, Supplement Figure S5). Although the precision of the inferred cluster was above 75% for 93% (88-93%) of the cases, 31% (6-39%) had a sensitivity score below 0.5, meaning they were classified with less than half of the cases from their reference clusters (Figure 4C). The discrepancies observed in this scenario are due to inconsistencies between the importation status distribution and the clustering of cases in the contact tracing investigations, as reference clusters that gathered several importations were split into different inferred clusters.

In scenario 3, we used different threshold *λ* to infer the importation status of cases. We tested *λ* = −15, *λ* = −10 (Absolute value), and *λ* = 95^th^ centile of all recorded log-likelhoods (Relative value). For each case *i*, if the log-likelihood *L_i_* was worse than *λ*, the connection between the case and its infector was removed and the case was considered imported. Firstly, using an absolute factor *λ* = −15, 586 (581-593) cases were classified as importations. 361 (355-369) of them were singletons. These numbers are much lower than the reference dataset that contains 737 clusters, and 539 singletons (Figure 5A, Supplement Figure S6). However, very few cases inferred as importations or singletons were not classified as such in the reference dataset (15 (10-22) misclassified importations, 4 (0-14) misclassified singletons), and the cluster size distribution for clusters including two cases and more was very similar to the reference one. The precision of the reconstructed cluster was very high (above 75% for 88% (85-93%) of cases) (Figure 5B). Overall, the algorithm was not able to accurately identify importations and singletons as the threshold was too low to eliminate some unrealistic connections, but the inferred larger clusters matched their reference counterparts.

**Figure 5:**
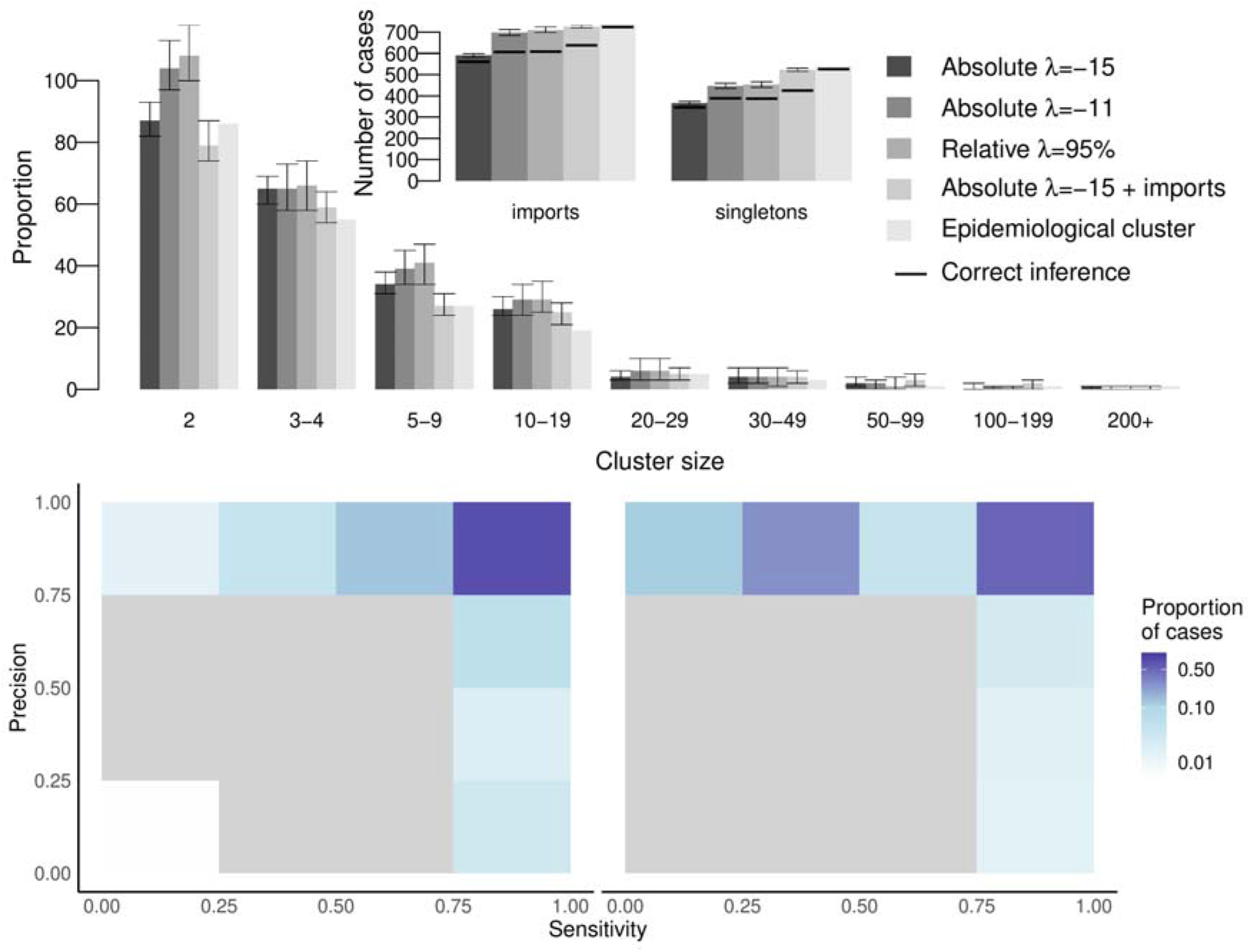
Description of transmission clusters generated with inferred importation status of cases. Panel A: Cluster size distribution for different value of threshold in the scenario 3 (sorted by shades of grey), compared to the reference clusters (lighgrey). Arrows represent the 95% credibility intervals of each estimate. Only clusters containing at least 2 cases are represented. Insert: Number of importations and number of isolated cases (singletons). For each scenario, the horizontal dark line represents the number of importations that are also importations in the reference clusters, same for singletons. Panel B: Heatmap representing the precision and sensitivity of the clusters for each case in scenario 3, with a 5% relative threshold, cases are classified in a category depending on the proportion of their reference cluster that were inferred in the same cluster. Panel C: Same when importation status is taken from the contact tracing investigations and inferred using a 5% relative threshold.

We then observed the impact of increasing *λ* on the inferred cluster size distribution. Runs obtained using an absolute threshold with *λ* = − 11 and 95% relative threshold yielded very similar results. The number of cases inferred as importations was higher than in previous runs, while all remaining links showed good connection between cases. The number of importations was closer to the reference dataset, and the number of singletons was greater than the reference. Nevertheless, the 11% (10-12%) of the inferred importations was not classified as importation in the reference clusters. Furthermore, the number of two-case chains was overestimated, and bigger clusters were likely to be split because of the removal of weaker connections. Therefore, increasing *λ* did not improve the cluster size distribution, as many importations in the reference clusters were not identified and the number of mismatches increased (Supplement Figures S7).

Finally, we combined prior information and inference of importation status to create a scenario where the importation status of only a proportion of the cases is known, because of disparities in the contact-tracing investigations. This scenario is relevant for a dataset combining different outbreaks scattered across a large area or a long period of time. Cases considered as importations in the contact tracing investigations were set as importations, and we inferred the importation status of the remaining cases. We used a low threshold, to remove the least likely transmission links (*λ* = − 15). Including prior information led to some misclassification of importation status due to the inconsistencies between the epidemiological importation status and the reference clusters. As in scenario 2, some cases were classified with only part of their reference clusters because clusters with several importations were split into different clusters. Indeed, the sensitivity score of 34% (7-51%) of cases was below 0.5. Nevertheless, the cluster size distribution observed in the simulation was the closest to the reference clusters. There were 725 (719-731) clusters, 89% of importations were also ancestors of reference clusters and the number of singletons matched the reference clusters (Figure 5A-C). The inferred clusters of 88% (86-94%) of the cases had a precision score of 1, showing they were clustered without any false positives. Despite discrepancies in several states (Massachusetts, Ohio), the cluster size distribution stratified by state showed good agreement with the reference clusters (Supplement Figure S8).

The conditional report ratio in the transmission chains *ρ* and the spatial parameters *a* and *b* was estimated in each scenario. The parameter estimates did not depend on the prior importation status distribution or the value of *λ*. *ρ* was consistently estimated above 90%, showing a low number of missing generations between cases (Supplement Figure S9). High values of *ρ* show that most of the reported cases could be connected without missing generations. This is not representative of the overall report ratio, which is usually much lower[51].

There was little variation in the estimates of the spatial parameters between the different scenarios. The population parameter *a* was estimated between 0.6 and 1 for every scenario, and the distance parameter b was between 0.08 and 0.12. In every scenario, more than 80% of the inferred transmission were between cases distant of less than 10km, and few long-distance transmissions were recorded (50-100km), hence although most of the reconstructed connections were between cases from the same county, the algorithm was able to identify clusters spreading over several counties or states (Supplement Figure S10).

We highlighted the added value of including the spatial distance between cases in the likelihood by comparing the cluster size distribution inferred by selecting certain components of *L_i_* (Supplement Figure S11). The credibility intervals were much wider when the distance between cases is not part of the likelihood, and the number of chains containing 2 to 10 cases was over estimated. The important impact of the spatial component of likelihood was also due to the widespread American territory, and could be lower in a different setting.

We used the ratio of the number of importations over the number of subsequent cases per state to evaluate the intensity of transmission in each state between 2001 and 2016 (Figure 6). The maps obtained in the scenario 1 (ideal scenario) or in scenario 3 (estimation of importation, with epidemiological importations and *λ* = − 15) were very similar. We only observed minor differences, for example in South Dakota and in Massachusetts, where the ratios were higher in scenario 3. The highest ratio (31.8 in scenario 1) was observed in Ohio, and is mostly due to a 383 case outbreak in 2014[32]. We observed major differences between the incidence map (Figure 2A) and the ratio per state. Indeed, although 403 cases were reported in California (highest number in the US), importations caused on average 1.32 subsequent cases in scenario 1 (1.60 in scenario 3), showing a high proportion of reported cases were inferred as importations.

**Figure 6:**
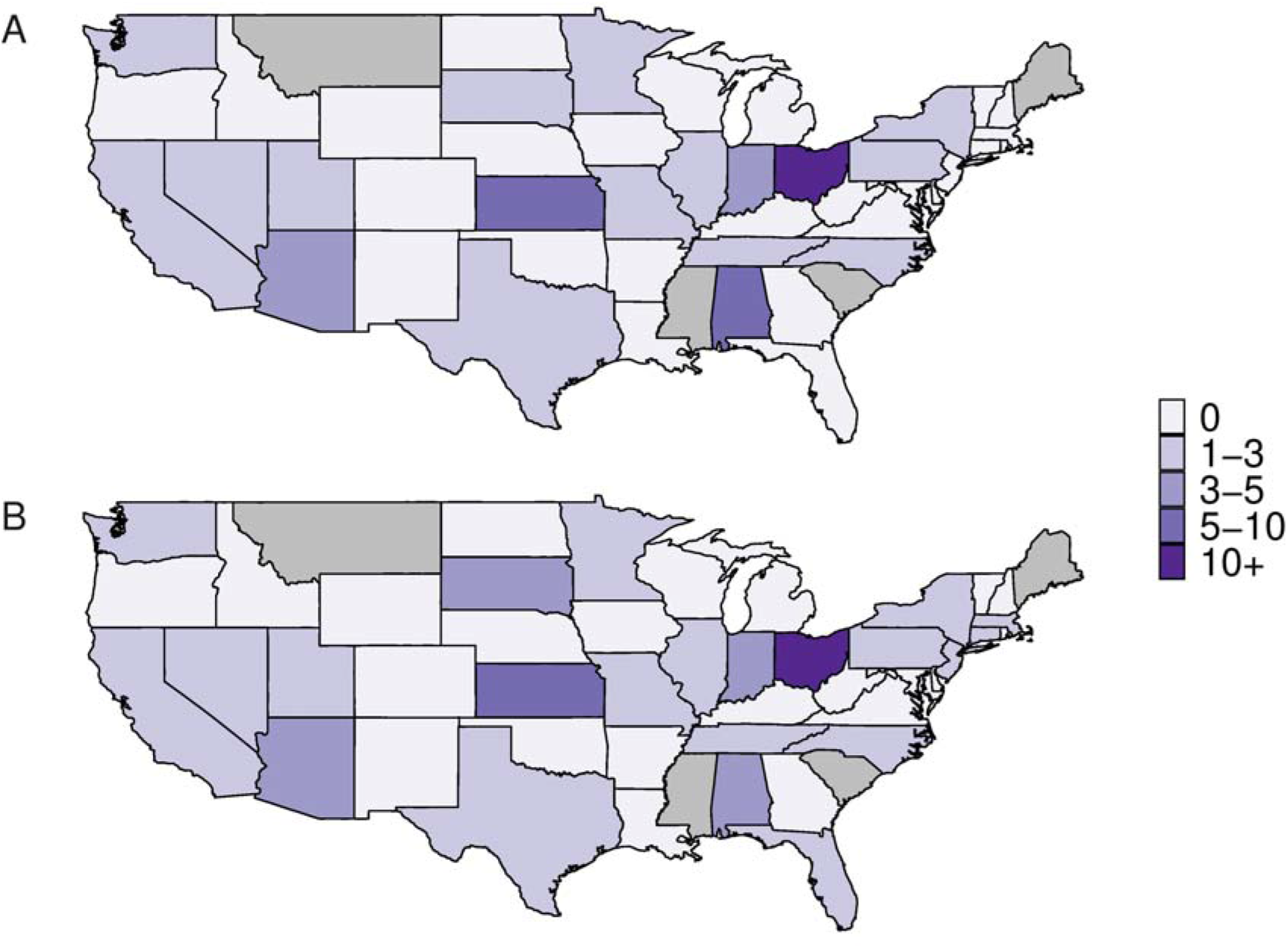
Ratio of the number of importations over the number of subsequent cases in each state in A/ Scenario 1 (Ideal importations) and B/ Scenario 3 with epidemiological importations and *λ =* −15. Grey states represent states that did not report any case.

Similarly, we used the inferred transmission chain to compute the inferred reproduction number in each state. According to the model, about 60% cases did not cause future transmission, and about 5% caused more than 5 subsequent cases (Supplement Figure S12). These numbers were consistent in each run. The geographical distribution of reproduction number was very similar to the importation - subsequent cases ratio (Supplement Figure S13).

## Discussion

We developed the R package *o2geosocial* to classify measles cases into transmission clusters and estimate their importation status using routinely collected surveillance data (genotype, age, onset date and location of the cases). As recently observed during the 2018-2019 measles outbreak in New York, delays in childhood vaccination, local susceptibility, and increased contacts can lead to large outbreaks following importations[52,53]. Therefore, we were interested in highlighting the effect of imported cases on communities and we focused on short distance transmission to identify areas where they repeatedly caused subsequent transmission chains. Although this is not predictive of future transmission, it highlights communities with potential for large transmission clusters.

We compared the inferred transmission clusters to the contact tracing investigations of 2,077 confirmed measles cases reported in the United States between 2001 and 2016. We were able to produce reliable estimates of known transmission clusters using epidemiological features with only few misclassifications. Estimating the importation status of cases without prior knowledge was challenging and caused uncertainty on the results. We tested different threshold to eliminate unlikely transmissions, and we were able to identify most of the imported cases. Nevertheless, if several cases were imported in the same region at a similar time, we could not find all of them without discarding valid transmission events, and increasing the number of false positives. When we used the importation status as defined in the contact tracing investigations without probabilistic inference (scenario 1 and 2), the reconstructed clusters were similar to the reference ones. Results were also conclusive when we combined prior information and importation inference. The reconstruction of transmission greatly depends on the epidemiological investigations to identify measles importations in a community.

We used the genotype to censor connections between cases when it was reported, as there can be only one reported genotype per transmission cluster. Using a simulated dataset (*toy_outbreak_long* in *o2geosocial)*, we explored the impact of increasing the proportion of genotyped cases on clustering and observed it could help identify the number of concurrent transmission trees when multiple genotypes are co-circulating. Moreover, we introduced a spatial component to the likelihood of connection between cases using an exponential gravity model. Previous studies showed this model was able to capture short distance dynamics better than other gravity models, and was easy to parametrise. Introducing the spatial component greatly improved the precision and the sensitivity of the reconstructed clusters (Supplement Figure S11), and the parameter estimates were robust in the different scenarios.

The final results on the clustering of the 2,077 cases using *o2geosocial* were obtained in 7 hours for each run of 100,000 iterations on a standard desktop computer (Intel Core i7, 3.20 GHz 6 cores), which is much faster than previous implementations of *outbreaker* and *outbreaker2*. With the addition of the pre-clustering step, whereby we reduced the number of potential infectors for each case, the algorithm ran faster. For smaller chains (50,000 iterations), 4 hours were needed to estimate the importation status and cluster the cases. The code for the package and the analysis developed in this project is shared on Github (*https://github.com/aJxsrobert/o2geosocial* and *alxsrobert/datapaperMO)*, with an illustrative toy dataset, and can be used to analyse recent outbreaks where contact-tracing investigations were not carried out.

Although the results obtained are promising, it should be noted that the dynamics of measles transmission in the United States are likely to be very specific to this location. Indeed, there were less than 700 annual cases between 2001 and 2016. These cases were scattered across a large area, which made the pre-clustering of cases very efficient as we focused on short-distance transmission. In smaller or more endemic settings, the number of potential infectors per cases after the pre-clustering step might be higher, which would increase the running time.

Furthermore, as the location of each case was deduced from the population centroid of counties, we assumed that the distance between cases from the same county was effectively zero. American counties are large and widespread geographical units that can include more than 1 million individuals. For future use of *o2geosocial*, more accurate information on the location of cases could improve cluster inference by identifying multiple importations in a given county. Because cases are reported by the state of residency, we had to ignore that cases may have been out of the reported county or state during their incubation and infectious period, which has been seen during some outbreaks, such as the 2015 “Disney outbreak” in California[54].

We did not include prior information on the local susceptibility of the different areas affected in *o2geosocial*, and these could be estimated using historical values of local coverage. However, protocols to estimate local vaccination coverage can differ in time and space and be difficult to compare, or unavailable at the local level. Furthermore, these estimates are cross-sectional in nature, and might not take into account catch-up vaccination campaigns, or immunity induced by previous outbreaks. Local seroprevalence surveys could identify pockets of susceptibles, but they have not been carried out on a subnational scale in most countries[55].

There has been no national quantitative analysis of age-specific contact patterns carried out in the United States, so we relied on a contact matrix between age-groups available for Great Britain from the POLYMOD study[36]. Nevertheless, little variation in the contact rates between age groups has been observed between European countries, and a previous projection of the social contact matrix in the United States yielded similar results[56]. POLYMOD data was probably the most reliable source of information we could use to deduce an estimate of the contact matrix in the United States.

## Conclusions

Heterogeneity in immunity can cause large outbreaks in countries with high national vaccine coverage, and identifying potential foyers of transmission in post-elimination settings is key for outbreak prevention and control. We have presented a method for estimating the cluster size distribution of past measles outbreaks from routinely collected surveillance data. We found that adding prior knowledge on the importation status of cases improved the inference of the transmission clusters. Although the method was able to identify a proportion of importations, epidemiological investigations on the history of travel and exposure reduced uncertainty on the clustering of cases. We believe these investigations are needed to produce reliable estimates of past transmission clusters. In lieu of the importation status, if multiple genotypes are co-circulating, increasing the proportion of genotyped cases could help discard potential connections and find imported cases. Even with limited information, this method was able to infer probabilistic transmission clusters in a fast and efficient way.

## Data Availability

The package we developed is publicly available on Github (https://github.com/alxsrobert/o2geosocial), along with the code used to analyse the data and generate the figures (https://github.com/alxsrobert/datapaperMO). Combinations of variables in the surveillance data used to validate this algorithm may contain sensitive personally identifiable health information which are subject to the Privacy Act and cannot be shared publicly. A toy dataset was attached to the o2geosocial package (in o2geosocial/data). The script analysis_generated_data.R in the datapaperMO repository generates toy datasets with different parameters (distance kernel, number of cases, reproduction numbers..) and can be used to re-run the model and test its performance.

https://github.com/alxsrobert/datapaperMO

## Acknowledgements

We acknowledge Thibaut Jombart for technical support and feedback on the analysis plan.

## Funding

AR was supported by the Medical Research Council (MR/N013638/1). SF was supported by a Wellcome Trust Senior Research Fellowship in Basic Biomedical Science (210758/Z/18/Z). AJK was supported by a Sir Henry Dale Fellowship jointly funded by the Wellcome Trust and the Royal Society (206250/Z/17/Z).

## Disclaimer

The findings and conclusions in this report are those of the authors and do not necessarily represent the official position of the Centers for Disease Control and Prevention, US Department of Health and Human Services.

## Author Contributions

AR, SF and AJK developed the method and the analysis plan. PAG and PP provided data for the study and gave feedback on the analysis plan. AR implemented the analysis, wrote the code and ran the model. AR interpreted the results, with contributions from SF, AJK and PAG. AR wrote the first draft and the supplementary material. AR, SF, AJK, PAG and PP contributed to the manuscript, all authors approved the final version.

## Notes

### Competing Interest Statement

The authors have declared no competing interest.

